# COVID-19 Vaccine Acceptance and Hesitancy in Cameroon: A Systematic Review and Meta-analysis

**DOI:** 10.1101/2024.12.12.24318938

**Authors:** Fabrice Zobel Lekeumo Cheuyem, Adidja Amani, Iyawa Clarisse Alma Nkodo, Lionel Bethold Keubou Boukeng, Michel Franck Edzamba, Ariane Nouko, Edwige Omona Guissana, Christelle Sandrine Ngos, Chabeja Achangwa, Christian Mouangue

**Author notes:** Corresponding author’s address: Fabrice Zobel Lekeumo Cheuyem, Department of Public Health, Faculty of Medicine & Biomedical Sciences, The University of Yaounde I, PO Box. 8526, Yaounde, Cameroon.

## Abstract

**Background:** The development of effective vaccines was a promising tool for ending the pandemic. However, the success of a vaccination programme depends heavily on achieving significant community acceptance. In Cameroon, numerous studies have investigated the level of acceptance, hesitancy and perception of COVID-19 vaccines, with mixed results. To provide a comprehensive understanding of these parameters, this meta-analysis aimed to estimate the pooled proportion of COVID-19 vaccine acceptance, hesitancy and perception in Cameroon.

**Methods:** A systematic search of online databases, including PubMed, Google Scholar, and ScienceDirect, was conducted to identify relevant research articles. This study followed the Preferred Reporting Items for Systematic Reviews and Meta-Analyses (PRISMA) guidelines. The extracted data were compiled in a Microsoft Excel spreadsheet and analyzed using R statistical software (version 4.3.3). The pooled proportion of COVID-19 vaccine acceptance, hesitancy, and perception was calculated using random effects meta-analysis. Funnel plots, Egger’s and Begg’s tests were used to assess publication bias.

**Results:** Of the 1243 records identified through the database search, 20 research articles were included in the systematic review and meta-analysis. The random-effects model showed that the that approximately 31.21% (95% CI: 23.49-38.94) of the Cameroonian population was willing to accept the COVID-19 vaccine. More than two-thirds of the population (68.49%; 95% CI: 60.65-76.34) were vaccine hesitant. Half of the participants (51.81%; 95% CI: 42.70-60.93), had a negative perception of the COVID-19 vaccine. The acceptance rate progressed from the first semester of 2021 (27.21%; 95% CI: 10.38-44.05) to the first semester of 2022 (45.56%; 95% CI: 25-66.12). The pooled vaccine acceptance rate was 29.29% (95% CI: 19.86-38.72) for the general population and 39.24% (95% CI: 22.84-55.64) for healthcare workers. The pooled vaccine hesitancy rate was 70.39% (95% CI: 61.30-79.80) for the general population and 57.42 % (95% CI: 4.05-71.80) for healthcare workers

**Conclusion:** Despite progress in vaccine acceptance, targeted interventions are still needed to address vaccine hesitancy in the country. Strategies such as improving access to accurate information, building trust in institutions, and strengthening community engagement are crucial to increasing COVID-19 vaccine uptake.

## Background

Coronavirus disease (COVID-19) is an infectious disease caused by the severe acute respiratory syndrome 2 (SARS-CoV-2) viruses [1]. The pandemic has resulted in an unprecedented burden on global healthcare systems and economies with a particular concern in sub-Saharan Africa [2]. Despite declining case numbers, periodic surges continue, resulting in more than 704 million confirmed cases and more than 7 million deaths worldwide as of 3 March 2024 [3].

In Cameroon, the first confirmed case of COVID-19 was reported on March 6, 2020. Following this first case, the government of Cameroon has implemented several preventive measures to limit the spread of the virus including vaccination [4]. The Cameroonian health authorities, represented by the National Immunization Technical Advisory Groups and the Scientific Advice for Public Health Emergencies, have approved four vaccines against SARS-CoV-2 for immunization against this disease [5].

Although vaccines against COVID-19 were developed, many factors compromise their acceptance and this become a public concern [6,7]. Vaccine hesitancy is one of the top ten global health problems. A distorted perception of disease risk, a lack of knowledge about vaccines, a fear of side effects, and the proliferation of misinformation and fake news are some of the key elements contributing to this public health problem [8]. Vaccine hesitancy negatively affects the vaccine uptake and hampers efforts to control the pandemic.

A study report found that despite the implementation of community engagement focused on education about vaccine efficacy, tolerability, and potential side effects, vaccine hesitancy was expected to persist in Cameroon between 2021 and 2022 [3].

Various studies have been conducted in Cameroon regarding the level of acceptance as well as hesitation and perception of COVID-19 vaccine. These levels vary between studies and the present meta-analysis was used to estimate the pooled proportion of these parameters in Cameroon.

## Methods

### Study Design

This study was conducted to assessed the proportion of vaccine acceptance and hesitancy in Cameroon. This systematic review was conducted in accordance with Preferred Reporting Items for Systematic Reviews and Meta-Analysis (PRISMA) guideline [9].

### Study Setting

Cameroon has an estimated population of about 28.6 million in 2023. The country covers a total area of 472,650 km², divided into ten administrative regions: Center, Littoral, Far North, North, Adamawa, North-West, South-West, West, East and South. Cameroon has two capitals: Yaoundé, located in the Center Region, serves as the country’s political capital, while Douala, located in the Littoral Region, is the economic city that drives the country’s growth [10].

### Eligibility Criteria

The review included all published cross-sectional studies reporting on COVID-19 vaccine acceptance, hesitancy and perception in Cameroon. Studies with unclear outcome variables were excluded from the review. In addition, duplicate articles were reviewed and removed from the study prior to data extraction. Only articles written in English language were included. There was no restriction on publication date as there was no previous systematic review and meta-analysis that investigated vaccine acceptance, hesitancy and perception in the country.

### Article Searching Strategy

Based on predetermined eligibility criteria, a comprehensive review of online literature sources was conducted. A systematic search of electronic databases such as PubMed, Google Scholar, and Science Direct was performed to identify published studies. The search strategy included analysis of the text contained in the title and abstract of each study. A combination of keywords and Medical Subject Headings (MeSH) terms was used, with Boolean logic operators (“AND” and “OR”) used to refine the search. The keywords and MeSH terms included “coronavirus OR COVID-19 AND vaccine AND acceptance OR willingness OR hesitation OR intention OR perception OR attitude AND Cameroon”. To ensure a comprehensive search, a manual search was conducted to identify additional published articles not indexed in electronic databases. To reach literature saturation, the reference lists of the indicated studies were also examined. The last search was conducted on November 15, 2024.

### Data Extraction

Data extraction was performed from all eligible articles. A predefined Microsoft Excel 2016 form was used to collect study characteristics including primary author, study year, region, study design, setting, study participant, sample size, reported vaccine acceptance, hesitancy, and poor perception. Two authors conducted a critical assessment of each article for relevance and quality. Disagreements among reviewers were resolved through discussion.

### Data Quality Assessment

The quality of the included studies was assesed using the Joanna Briggs Institute quality assessment tool for prevalence studies [11]. Nine parameters were employed to assess the risk of bias for each study. Such parameters include appropriateness of sampling frame, suitable sampling technique, adequate sample size, description of study subjects and setting, sufficient data analysis, use of valid methods for identified conditions, valid measurement for all participants, use of appropriate statistical analysis, and adequate response rate (≥60%). Each parameter was scored as 1 (yes) or 0 (no or unclear). The risk of bias was categorized as low (5-9), moderate (3-4), or high (0-2).

### Outcome of Measurement

The primary outcomes of this study were COVID-19 vaccine acceptance and hesitancy. The other outcome variable included the negative perception this vaccine. These proportions were calculated by dividing the number of participants who indicated willingness to receive the vaccine, hesitancy to receive the vaccine, or negative perception of the vaccine by the total number of participants who responded to the question.

### Operational Definition

Vaccine acceptance refers to intention or willingness to receive the vaccine and not actual administration (uptake) of the vaccine itself. Vaccine hesitancy is the refusal of vaccines or a delay in acceptance despite the availability of immunization services [4]. Negative or poor perception of the COVID-19 vaccine refers to a negative attitude toward the benefits of the COVID-19 vaccine. It includes cognitive and assessment of the vaccine as undesirable, untrustworthy, dangerous or ineffective, resulting in reluctance or refusal to receive the vaccine.

### Statistical Analysis

Data were analyzed using R Statistics version 4.3.3. Heterogeneity between studies was assessed using the *I^2^* test statistic. Three categories were used to classify the degree of heterogeneity (*I^2^*index). These categories were either low (<25%), moderate (25-75%), or high (>75%). Subgroup analysis was performed for study year, region, setting, and type of participants enrolled. The random effects model was preferred when significant heterogeneity between studies was observed for the pooled estimates of vaccine acceptance, hesitancy, and perception.

Meta-regression was performed to investigate whether study characteristics could explain the variability in results across studies. The examined study characteristics included the year the study was conducted (2020, 2021, 2022 or 2023), region (studies targeting participants from all ten Regions (nationwide) of the country or less Regions (multicenter) or from a specific Region), setting (participants enrolled from hospital or from other settings including online-and community-recruited participants), sample size (<1000 and ≥1000), study participant (general or other that includes healthcare workers and/or students). Only study variables with meaningful and practical categories were considered. multivariable meta-regression model was used to assess whether vaccination acceptance, hesitancy, and perception varied according to the selected study variables. A *p*-value<0.05 was considered statistically significant.

### Publication Bias and Sensitivity test

Publication bias was assessed visually using the funnel plot. A funnel plot displaying symmetrical, large, and inverted shapes, suggested the absence of publication bias. Furthermore, Egger’s and Begg’s tests were used to statistically examine the funnel plot asymmetry, with a significance level of *p*<0.05. The sensitivity test was done by excluding a single individual study from the analysis at a time to explore the robustness of the findings.

## Results

A total of 1243 records were identified after an extensive literature search. After removing 135 duplicate studies, 1231 records remained. After review, 1208 reports were excluded based on their titles and abstracts. The content of the remaining 23 records was then assessed for eligibility and a total of 20 study reports met the eligibility criteria and were included in this systematic review and meta-analysis (Fig. 1).

### Selection of Studies

**Fig. 1.**
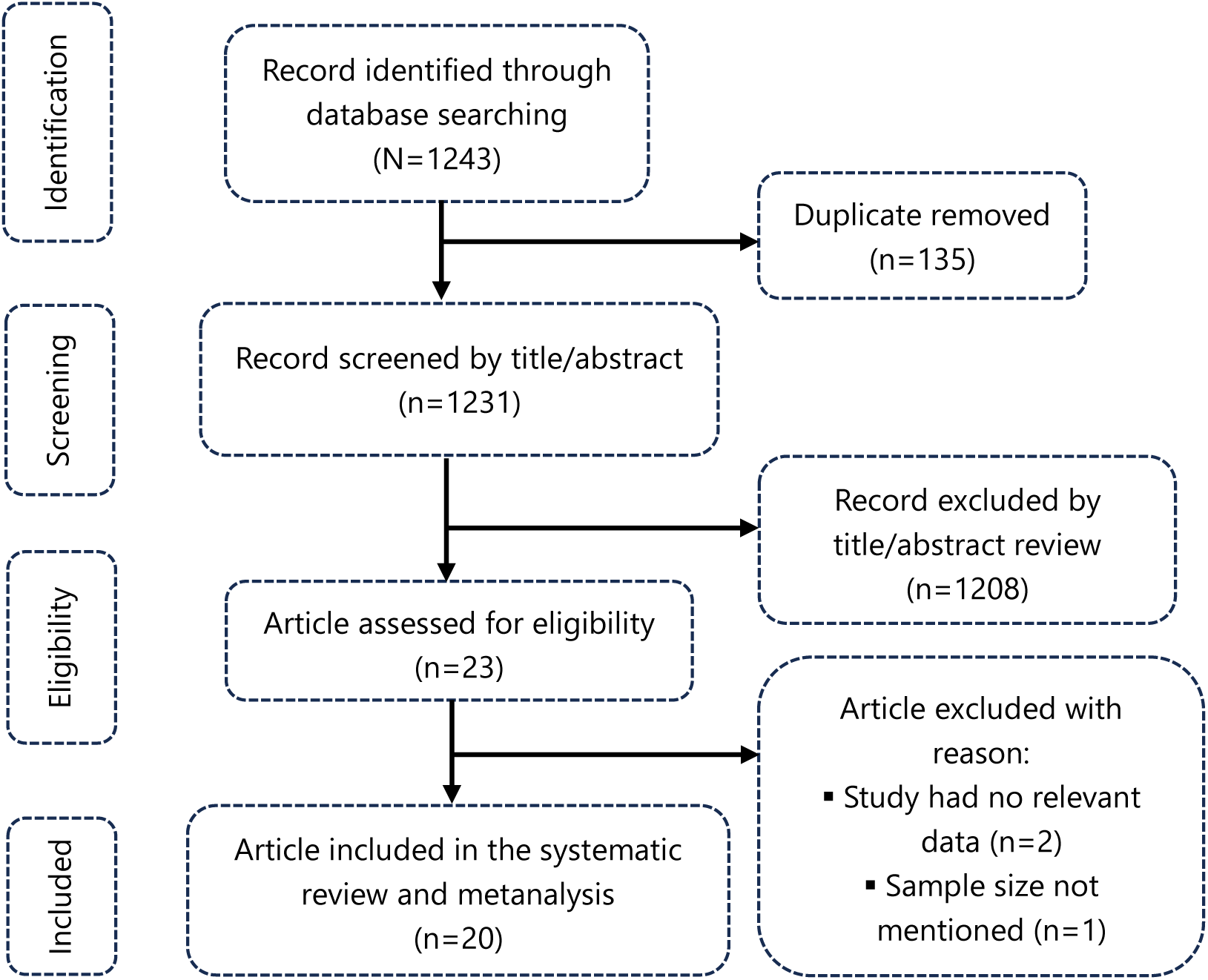
PRISMA diagram flow of studies included in the metanalysis.

### Summary of Studies Included

A comprehensive analysis included 20 studies with a cumulative sample size of 28,355 participants. These studies, conducted between 2021 and 2023, primarily involved the general population in different regions of the country and provided estimates of COVID-19 vaccine acceptance, hesitancy, and perception (Table 1).

**Table 1.** Characteristic of studies assessing adherence to COVID-19 vaccine in Cameroon, 2020-2023.

| Author | Study year | Region | Setting | Study population | Design | Sample size | Sampling | Risk of bias |
| --- | --- | --- | --- | --- | --- | --- | --- | --- |
| Abongwa *et al.* [12] | 2021 | North-west | Community | General population | CS | 2531 | Non-probabilistic | Low |
| Aka *et al.* [13] | 2022-2023 | Centre | Hospital | HCW | CS | 510 | Probabilistic | Low |
| Akwa *et al.* [14] | 2021 | Nationwide | Online | General population | CS | 1750 | Non-probabilistic | Low |
| Amani *et al.* [4] | 2021 | Nationwide | Online | General population | CS | 150 | Non-probabilistic | Low |
| Ambe *et al.* [15] | 2022 | Sout-west | Hospital | Nurse | CS | 197 | Probabilistic | Low |
| Aseneh *et al.* [16] | 2021 | National | Online | General population | CS | 341 | Non-probabilistic | Low |
| Baecher *et al.* [17] | 2022 | North-west<br>South-west<br>Littoral<br>Centre | Hospital | HCW | CS | 825 | Non-probabilistic | High |
| Chefor *et al.* [18] | 2021 | Centre | Hospital | HCW | CS | 247 | Non-probabilistic | Low |
| Cho *et al.* [19] | 2021 | West Littoral<br>Centre | Online | General population | CS | 665 | Non-probabilistic | Low |
| Dinga *et al.* [20] | 2020 | Nationwide | Online and<br>Community | General population | CS | 2512 | Non-probabilistic | Low |
| Dinga *et al.* [21] | 2022 | Nationwide | Online and<br>Community | General population | CS | 6732 | Non-probabilistic | Low |
| Djuikoue *et al.* [22] | 2021 | Littoral<br>West | Community | General population | CS | 1053 | Probabilistic | Low |
| Elit *et al.* [23] | 2022 | North-west | Community | General population | CS | 31 | Non-probabilistic | Moderate |
| Gunawardhana *et al.* [24] | 2021 | Nationwide | Hospital | General population | CS | 835 | Non-probabilistic | Low |
| Ajonina-Ekoti *et al.* [25] | 2021 | Littoral | Online and<br>University | Student | CS | 591 | Non-probabilistic | Low |
| Ngasa *et al.* [26] | 2021 | Nationwide | Online | HCW and<br>Student | CS | 371 | Non-probabilistic | Low |
| Tambo *et al.* [27] | 2021 | Centre | Community | General population | CS | 1522 | Probabilistic | Low |
| Tchiasso *et al.* [3] | 2021-2022 | Nationwide | Community | General population | CS | 6567 | Probabilistic | Low |
| Tetsatsi *et al.* [28] | 2022 | West | Community | General population | CS | 520 | Non-probabilistic | Low |
| Ukah *et al.* [29] | 2022 | Sout-west | Hospital | HCW | CS | 405 | Probabilistic | Low |

CS: Cross section study; HCW: Healthcare worker

### Adherence to COVID-19 Vaccine

The random effects model showed that approximately 31.21% (95% CI: 23.49-38.94) of the Cameroonian population was willing to accept the COVID-19 vaccine, with significant heterogeneity observed (*I^2^*=99.8%; *p*<0.001). Conversely, more than two-thirds of the population (68.49%; 95% CI: 60.65-76.34), were vaccine hesitant with a high heterogeneity of studies assessed (*I^2^*=99.3%; *p*<0.001). In addition, half of the participants (51.81%; 95% CI: 42.70-60.93), had a negative perception of the COVID-19 vaccine, with significant heterogeneity (*I^2^*=97.2%; *p*<0.001) (Fig. 2).

**Fig. 2.**
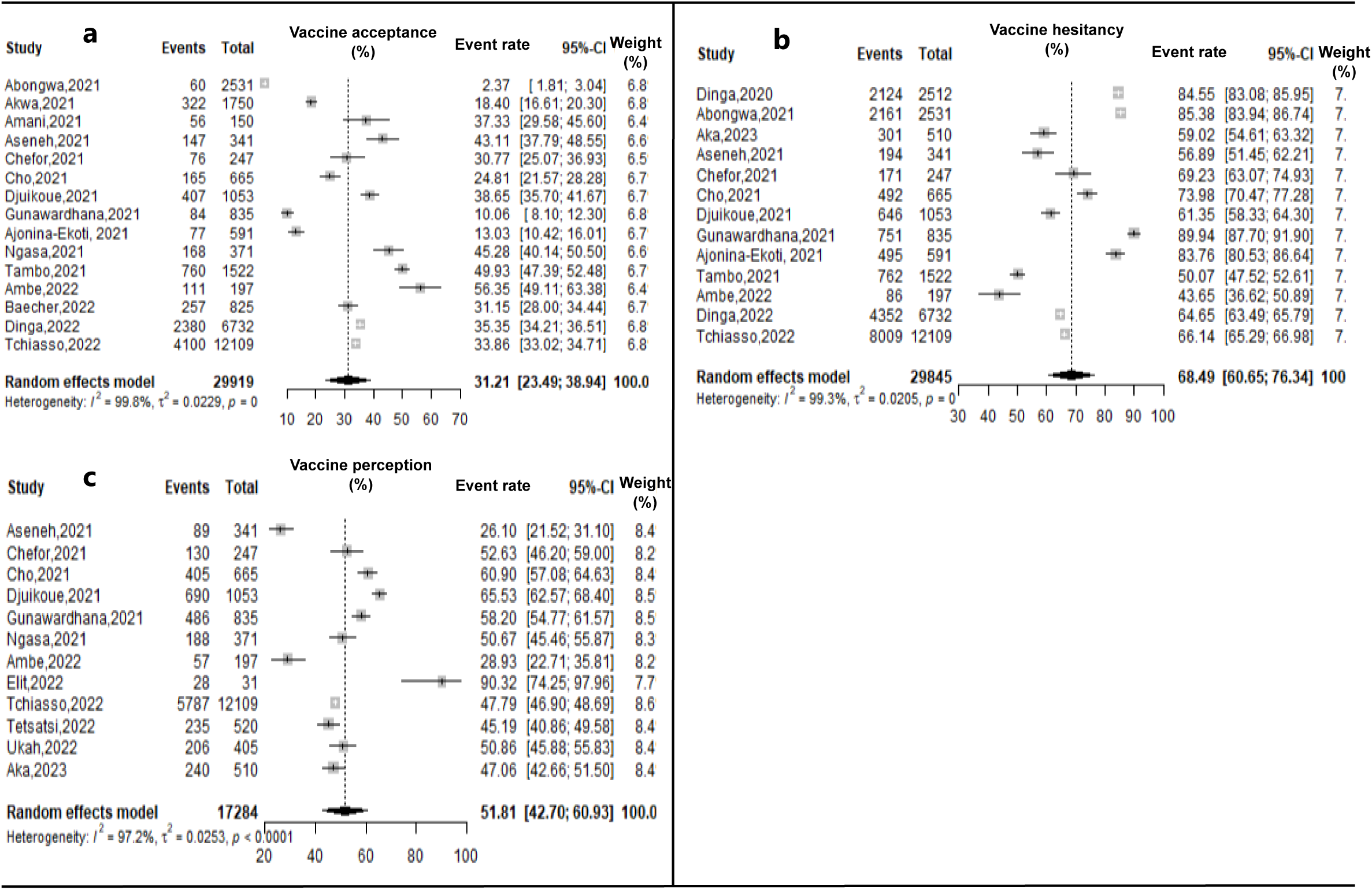
Forest plot displaying the COVID-19 vaccine acceptance (a), hesitancy (b) and negative perception rates (c) in Cameroon, 2020-2023.

### Subgroup Analysis

The lowest estimates of the willingness to receive COVID-19 vaccine were observed in the North-west Region (2.37 %; 95% CI: 1.91-3.04) and among students (13.03%; 95% CI: 45.73-83.06). The acceptance rate progressed from the first semester of 2021 (27.21%; 95% CI: 10.38-44.05) to the first semester of 2022 (45.56%; 95% CI: 25-66.12). The pooled vaccine acceptance rate was 29.29% (95% CI: 19.86-38.72) for the general population and 39.24% (95% CI: 22.84-55.64) for healthcare workers (Fig. 3).

**Fig. 3.**
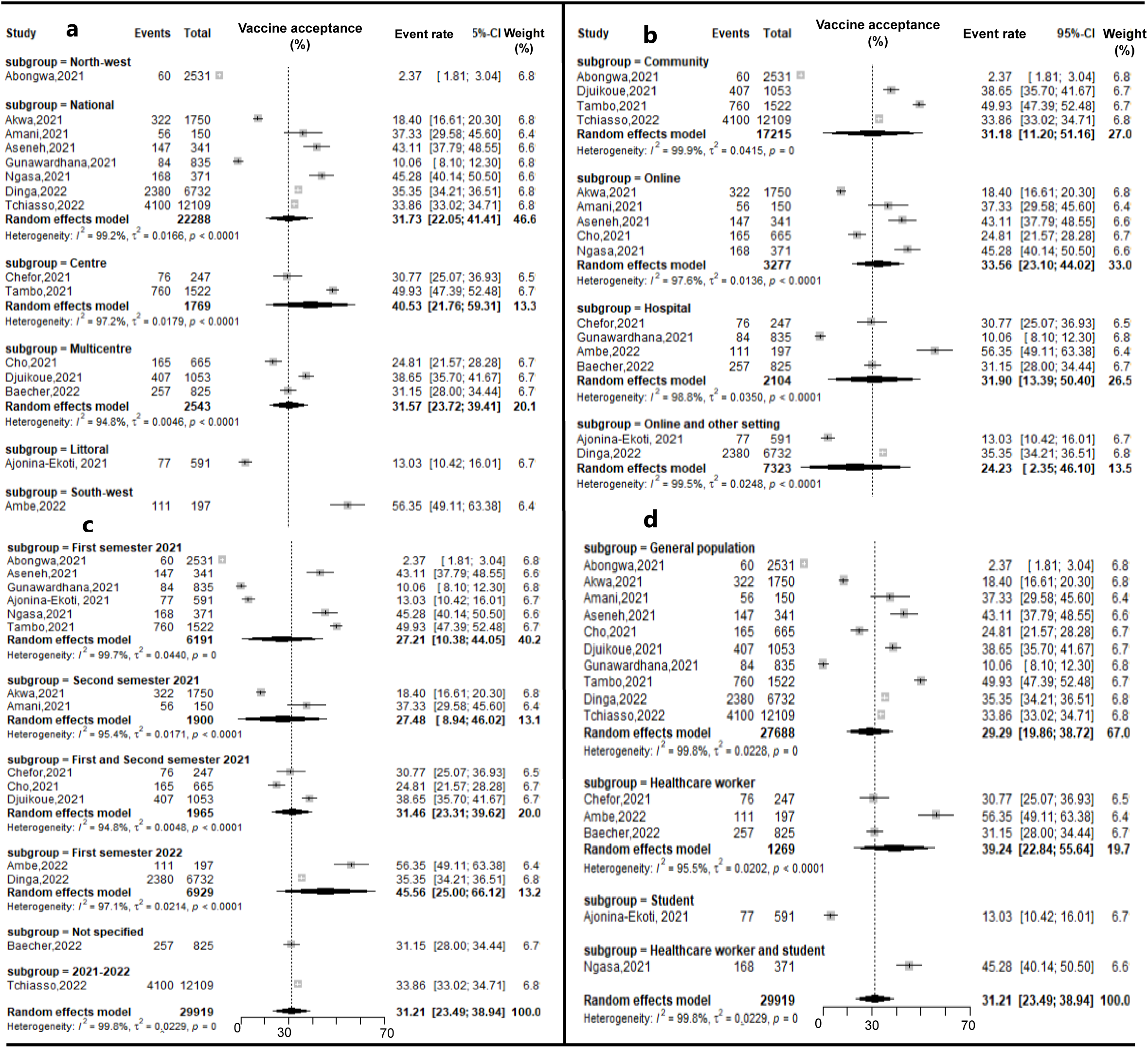
Subgroup analysis reporting the COVID-19 vaccine acceptance rate in Cameroon, 2021-2023 *(a: by Region; b: by setting; c: by study period and d: by type of participants)*

The highest hesitancy rate was observed in the half of 2021 with a pooled estimate of 75.96% (95% CI: 63.24-88.68) and in studies conducted both online and in other settings (77.62%; 95% CI: 64.83-90.41). The pooled vaccine hesitancy rate was 70.39% (95% CI: 61.30-79.80) for the general population and 57.42 % (95% CI: 4.05-71.80) for healthcare workers (Fig. 4).

**Fig. 4.**
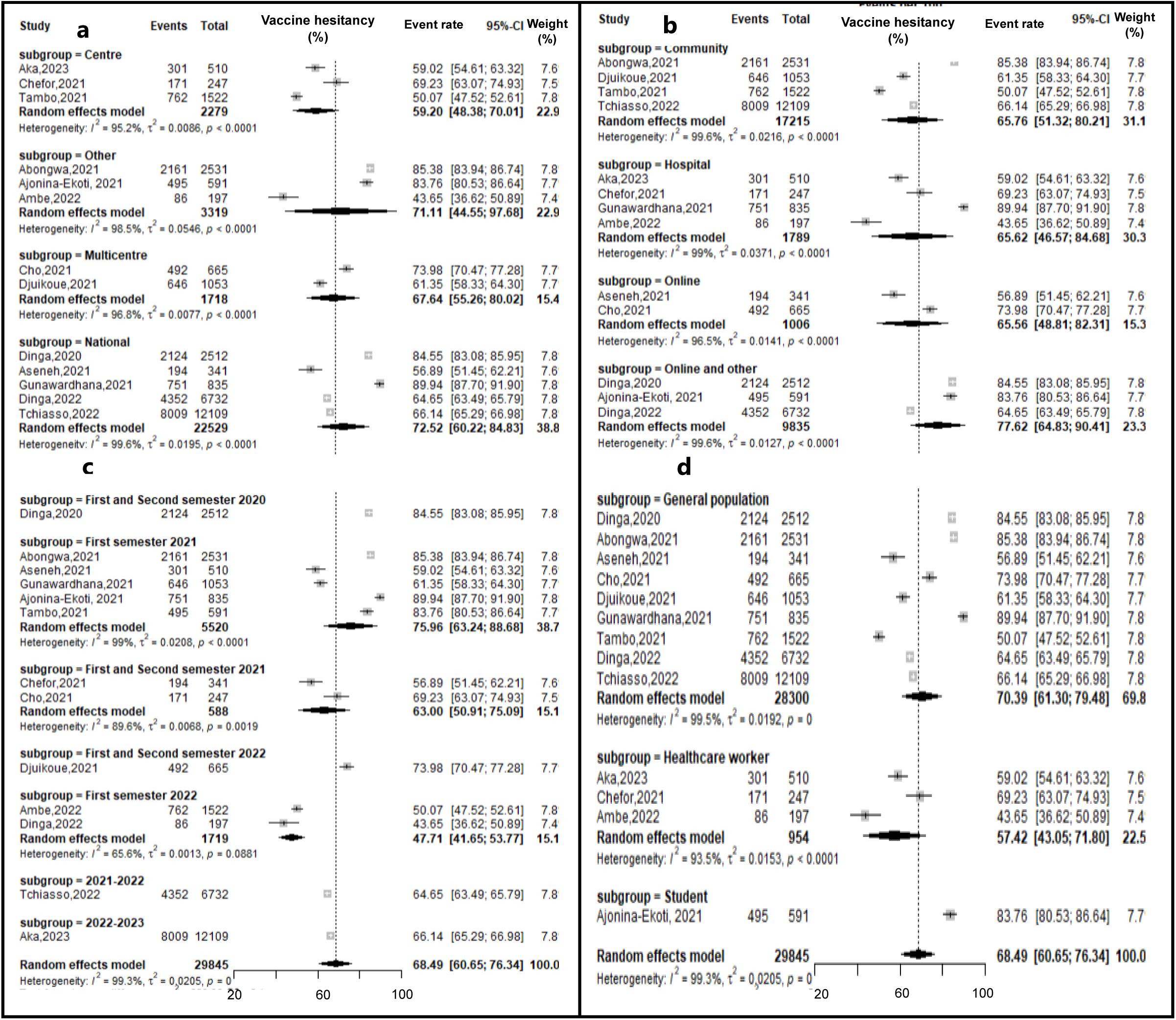
Forest plot showing the subgroup analysis of the COVID-19 vaccine hesitancy rate in Cameroon, 2020-2023 *(a: by Region; b: by setting; c: by study period and d: by type of participants)*

Poor perception of COVID-19 vaccine was mainly observed in studies conducted in other setting (North-west and West Regions) (67.50%; 95% CI: 23.27-100), in multicenter studies (63.37%; 95% CI: 58.85-67.89), and among participant from community (61.79%; 95% CI: 41.86-81.72). The negative perception rate was 56.01% (95% CI: 41.63-70.40) in the general population and 44.96% (95% CI: 34.48-55.44) among healthcare workers (Fig. 5).

**Fig. 5.**
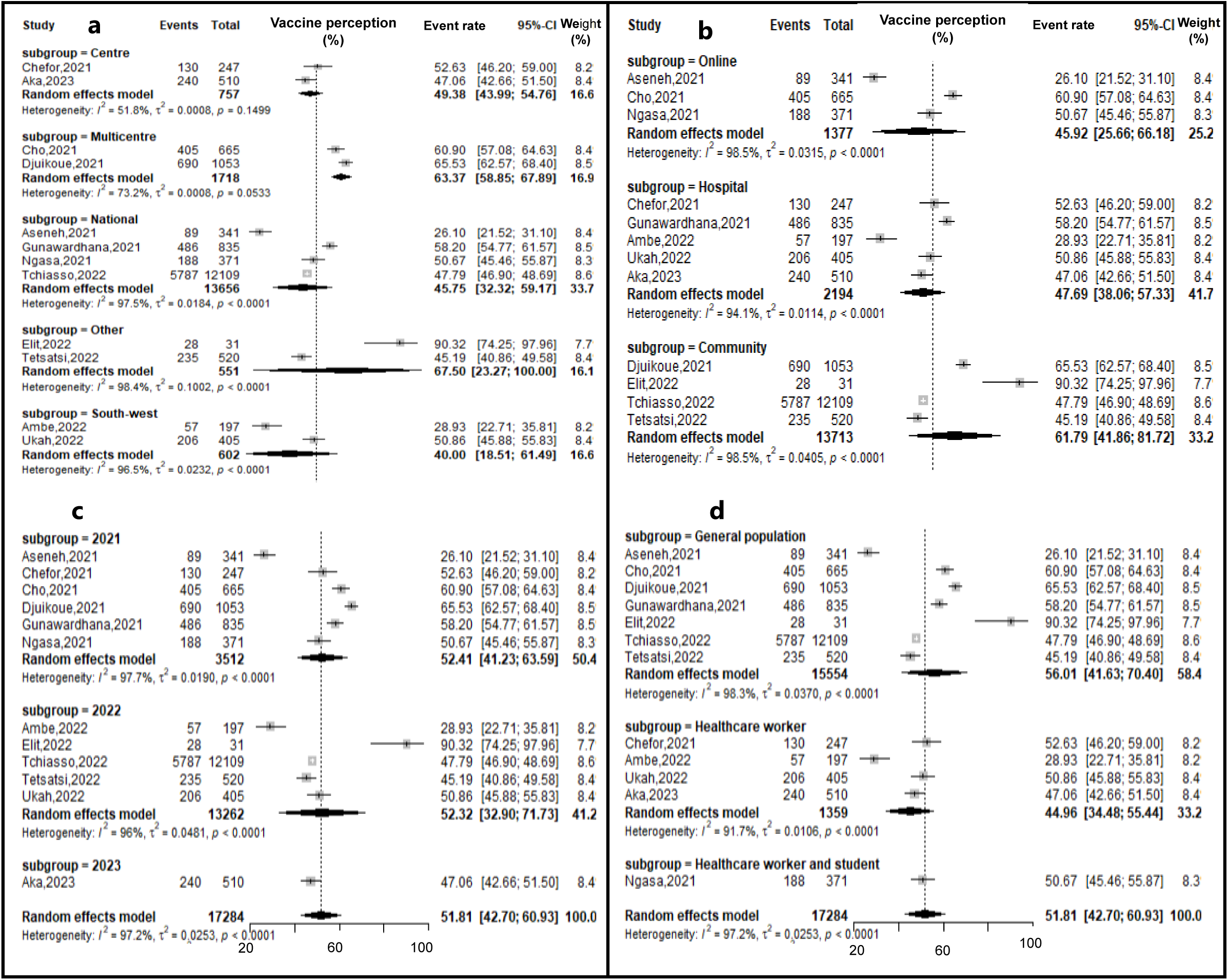
Negative perception subgroup estimates of the COVID-19 vaccine in Cameroon, 2021-2023 *(a: by Region; b: by setting; c: by study period and d: by type of participants)*

### Meta-Regression Analysis

The meta-regression analysis showed that the sampling method used had a significant effect on the heterogeneity of willingness to accept the COVID-19 vaccine (*p*=0.014). Surveys conducted before or during the introduction of the COVID-19 vaccine in Cameroon (2020-2021) revealed a significantly higher rate of vaccine hesitancy than those conducted in the following years (2022-2023) (*p*<0.001). In addition, vaccine hesitancy was higher in studies conducted in other settings (online or community) compared to those conducted in hospital settings (*p*<0.001) (Table 2).

**Table 2.** Multivariate metanalysis of COVID-19 vaccine acceptance, hesitancy and perception in Cameroon healthcare workers in Cameroon, 2020-2023.

| COVID-19 vaccine estimate | Category | Moderator | Adjusted coefficient ($\beta$) | p-value |
| --- | --- | --- | --- | --- |
| **Acceptance** | Study year | 2021 vs 2022 | -0.3965 | 0.553 |
|  | Setting | Other<sup>1</sup> vs Hospital | 0.6130 | 0.396 |
|  | Region | Subnational<sup>2</sup> vs Nationwide | -0.6440 | 0.268 |
|  | Sampling | Non-probabilistic vs Probabilistic | -1.6216 | **0.014** |
| | Sample size | <1000 vs $\geq 1000$ | 1.0653 | 0.1525 |
|  | Participant | Other<sup>3</sup> vs General population | 0.4229 | 0.570 |
| **Hesitancy** | Study year | 2020-2021 vs 2022-2023 | 1.2502 | **<0.001** |
|  | Setting | Other<sup>1</sup> vs Hospital | 1.4673 | **<0.001** |
|  | Region | Subnational<sup>2</sup> vs Nationwide | 0.1790 | 0.593 |
|  | Sampling | Non-probabilistic vs Probabilistic | 0.2866 | 0.342 |
| | Sample size | <1000 vs $\geq 1000$ | -0.9047 | **<0.001** |
|  | Participant | Other<sup>3</sup> vs General population | 0.9696 | **0.017** |
| **Negative perception** | Study year | 2021 vs 2022+ | 0.31143 | 0.6539 |
|  | Setting | Other<sup>1</sup> vs Hospital | -0.2460 | 0.750 |
|  | Region | Subnational<sup>2</sup> vs Nationwide | 0.7020 | 0.228 |
|  | Sampling | Non-probabilistic vs Probabilistic | 1.1791 | 0.323 |
| | Sample size | <1000 vs $\geq 1000$ | -1.2916 | 0.363 |
|  | Participant | Other<sup>3</sup> vs General population | -0.0620 | 0.936 |

<sup>1</sup>Online and/or within Community; <sup>2</sup>Multicenter and/or specific Region; <sup>3</sup>Healthcare workers and/or students

### Publication Bias and Sensitivity test Analysis

To assess publication bias, a traditional funnel plot was used, which showed asymmetry, indicating potential publication bias. For further investigation, Egger’s linear regression and Begg’s rank correlation tests were performed. Contrary to the visual impression of the funnel plot, the tests indicated no statistically significant publication bias for studies used to assessed COVID-19 vaccine acceptance (*p*=0.257), vaccine hesitancy (*p*=0.435) and perception (*p*=0.611) (Fig. 6).

**Fig. 6.**
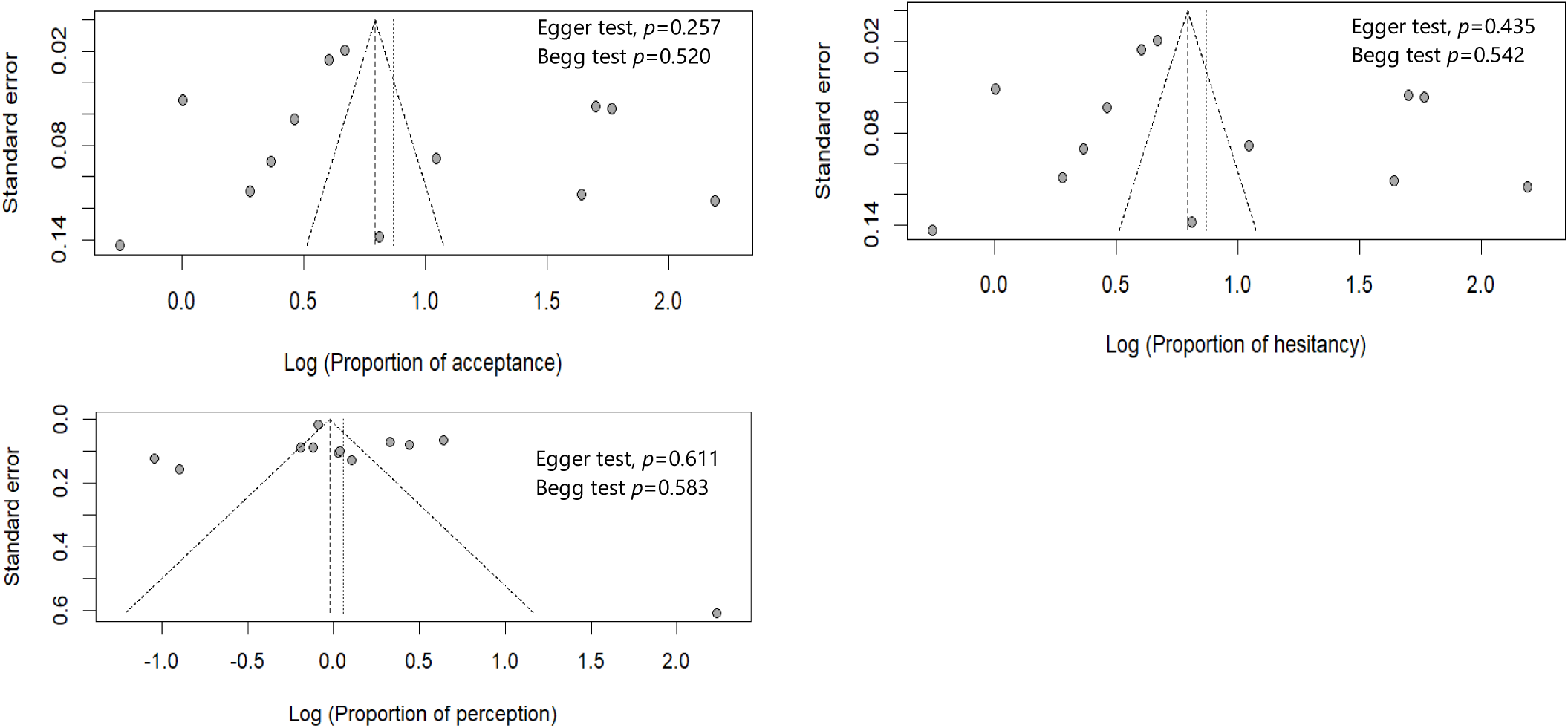
Funnel plot with pseudo 95% confidence limits and the resulting Egger’s and Begg’s tests of studies included

A sensitivity analysis was also performed to assess the impact of individual studies and outliers on the overall results. This analysis showed that no single study had a significant impact on the overall results (Supplemental tables 1,2 and 3).

## Discussion

The results of the study reveal a worrying trend in Cameroon, where approximately 31.21% of the population are willing to accept the COVID-19 vaccine, while a significant majority (68.49%) are hesitant to accept the vaccine. An almost similar trend was observed in a study conducted among large rural, underserved and minority populations in Alabama, USA [30]. This vaccine acceptance rate was lower than that observed in several meta-analyses conducted in Ethiopia and worldwide [1,6,21,31]. The high rate of vaccine hesitancy in the country could explain this acceptance rate. Vaccine hesitancy is a major concern as it may hinder efforts to control the spread of COVID-19. This finding corroborates results from studies conducted in the USA, where certain ethnic groups and underserved communities appeared to be more reluctant to be vaccinated against COVID-19 [32,33]. A much lower proportion of COVID-19 vaccine hesitancy was observed in a global meta-analysis conducted in 2023 [7].

In addition, about half of the participants (51.81%) have a negative perception of the vaccine. This negative perception may be due to various factors, such as misinformation, fear of side effects, or mistrust of the vaccine’s efficacy. This also explains the high hesitancy rate observed in the Cameroonian population.

The study found a significant increase in COVID-19 vaccine acceptance in Cameroon from the first semester of 2021 to the first semester of 2022. The acceptance rate increased from 27.21% in 2021 to 45.56% in 2022. This could be explained by the Ministry of Health’s community engagement efforts to sensitize the population with the aim of breaking down barriers to acceptance of the COVID-19 vaccine [3]. A natural process could also explain this progress, as humans are known to integrate new concepts over time. Our results are consistent with the findings of a global meta-analysis, where a significant increase in acceptance was observed from 2020 to 2021 [6,31].

The pooled vaccine hesitancy rate was 70.39% for the general population and 57.42% for healthcare workers. This result corroborates findings from a global meta-analysis where higher hesitancy was observed in the general population compared with a specific group including healthcare workers [31]. The general population may have limited access to credible sources of information about COVID-19 vaccines, making them more susceptible to misinformation and myths. In addition, the general population may be more skeptical of institutions such as the government, pharmaceutical companies, and healthcare systems, which may influence their attitudes towards vaccination. This lack of access to accurate information and mistrust can lead to confusion, suspicion, and ultimately, vaccine hesitancy.

### Limitations

Absence of hesitancy rate and a specific variable to assess poor perception of covid-19 rate in certain studies have reduced the number of studies included and should be considered in interpreting the result related to vaccine perception. Although the study checked for publication bias using funnel plots and Egger tests, it is possible that some relevant studies may not have been published or were not accessible. The study relied on self-reported data, which may be subject to social desirability bias, leading to overreporting of vaccine acceptance and underreporting of vaccine hesitancy.

### Conclusions

The study’s findings reveal a concerning trend of high vaccine hesitancy rates among the Cameroonian population. Willingness to receive the vaccine was lower than the global trend. The study also found a significant increase in COVID-19 vaccine acceptance rates in the country from 2021 to 2022. The results suggest that the general population in Cameroon is more reluctant to be vaccinated with COVID-19 than healthcare workers. The results of the study highlight the need for targeted interventions to address vaccine hesitancy in Cameroon. Efforts to improve access to accurate information, build trust in institutions and strengthen community engagement are critical to increasing acceptance of vaccine against COVID-19 or future emerging or re-emerging diseases.

## Data Availability

Data supporting this study are available in the reference. All data generated or analyzed during this study are included in this published article and supplemental material.

## Abbreviations

COVID-19: New Coronavirus Disease
CS: Cross Sectional Study
HCW: Healthcare Worker
MeSH: Medical Subject Headings
PRISMA: Preferred Reporting Items for Systematic Reviews and Meta-Analysis
SARS-CoV-2: Severe Acute Respiratory Syndrome 2

## Declarations

### Author contributions

FZLC conceived the original idea of the study. FZLC and MFE conducted the literature search. FZLC, MFE selected the studies, extracted the relevant information, and synthesized the data. FZLC performed the analyses and wrote the first draft of the manuscript. All authors critically reviewed and revised successive drafts of the manuscript. All authors read and approved the final manuscript.

### Ethical Approval Statement

Not applicable

### Consent for publication

Not applicable.

### Availability of data and materials

Data supporting this systematic review are available in the reference. All data generated or analyzed during this study are included in this published article.

